# Five Years, Two Mass Triple-Drug Administrations, and Ongoing Transmission: Using Mosquito and Human Indicators to Evaluate the Impact of Lymphatic Filariasis Interventions in Samoa

**DOI:** 10.64898/2025.12.24.25342955

**Authors:** Helen J. Mayfield, Angus McLure, Lisa Rigby, Brady McPherson, Richard Bradbury, Shannon Hedtke, Rhys Izuagbe, Jo Kizu, Donna MacKenzie, Elina Panahi, Beatris Mario Martin, Benn Sartorius, Asrhyella Seeler, Selina Ward, Robert Thomsen, Satupaitea Viali, Patricia M. Graves, Colleen L. Lau

## Abstract

**Background:** For lymphatic filariasis (LF) elimination, the World Health Organization recommends multiple rounds of mass drug administration (MDA). While LF antigen (Ag) is routinely used to monitor progress, recent evidence suggests more time-sensitive indicators are needed during the immediate post-MDA period. In Samoa, triple-drug MDA was distributed in 2018 and 2023. This study aimed to evaluate the impact of Samoa’s second round of triple-drug MDA on human and mosquito-based indicators.

**Methodology:** Surveys were conducted in eight primary sampling units (PSUs) in 2019 (7–9 months after the 2018 MDA), 2023 (4.5 years after the 2018 MDA) and 2024 (10 months after the 2023 MDA). Participants aged ≥5 years from randomly selected households were tested for Ag and microfilariae (Mf). For molecular xenomonitoring (MX), mosquitoes were caught using BG-Sentinel traps at households, sorted into pools by species, and tested for filarial DNA using quantitative polymerase chain reaction (qPCR).

**Results and key findings:** In 2024, Ag prevalence was 10.3% (95% CI:7.3-14.5) vs 9.9% (95% CI: 3.5-21.0) in 2023 and 9.8% (95% CI:5.6-15.5) in 2019. Mf prevalence was 3.1% (95% CI: 1.8-5.5) in 2024 vs 5.1% (95% CI:1.3-12.4) in 2023 and 3.1% (95% CI:1.3-5.9) in 2019. Odds ratios (OR) of a positive test in 2024 vs 2023 showed no decrease for Ag (OR 1.0; 95% CI:0.7-1.6), but a potential reduction in the proportion of Ag-positive participants who were Mf-positive (OR 0.6; 95% CI: 0.2-1.4) and a significant reduction in the prevalence of qPCR-positive mosquitoes (OR 0.4; 95%CrI: 0.2-0.7). From 2019-2024, there were reductions in prevalence of qPCR-positive *Aedes* spp. (OR 0.5; 95% CrI:0.3-1.0) and *Aedes polynesiensis* (OR 0.4; 95% CrI: 0.2-0.8).

**Conclusions:** In Samoa, LF transmission continues despite two rounds of triple-drug MDA five years apart. Mosquito indicators can provide a more sensitive measure of MDA impact in the immediate post-intervention period and complement human indicators for long-term surveillance.

**Author summary:** Lymphatic filariasis is a mosquito-borne disease that can cause severe long-term disability. To eliminate this neglected tropical disease as a public health problem, the World Health Organization (WHO) recommends multiple rounds of mass drug administration (MDA) 12 months apart. Despite antigen (Ag) being the standard indicator for evaluating MDA impact, more time sensitive indicators, such as measuring microfilariae (Mf) in the blood or testing mosquitoes for filarial DNA (molecular xenomonitoring) may provide a more reliable assessment in the months following the intervention. Samoa distributed two rounds of triple-drug MDA in 2018 and 2023. We evaluated the impact of the second round on human and mosquito-based infection indicators including Ag, Mf, and qPCR-positive mosquitoes.

We surveyed eight villages in Samoa in 2019 (7–9 months after the 2018 MDA), 2023 (4.5 years after the 2018 MDA) and 2024 (10 months after the 2023 MDA). The two rounds of MDA were insufficient to break the LF transmission cycle. Results from 2024 showed no decrease in Ag prevalence from either 2019 or 2023, although there was potentially a decrease in Mf prevalence. Molecular xenomonitoring (MX) provided an earlier signal of MDA impact than Ag, with a decrease in the prevalence of qPCR-positive mosquitoes observed within 10 months of the 2023 MDA.

## Introduction

Lymphatic filariasis (LF) is a mosquito-borne neglected tropical disease (NTD) that can cause debilitating lymphedema and scrotal hydroceles [1]. The disease is caused by infection from one of three parasites: *Wuchereria bancrofti*, *Brugia malayi,* and *Brugia timori*. The mosquito species that act as the vector vary from country to country and include species from the genera *Culex, Aedes, Anopheles,* and *Mansonia* [2]. Adult parasites reside and reproduce in the human lymphatic system, from which microfilariae (Mf) make their way into the circulating bloodstream, where they can be ingested by blood-feeding mosquitoes. Mf mature into their larval stages inside the thoracic muscles of mosquitoes before being transmitted to humans through a bite [3].

Despite decades of elimination efforts as part of the Global Programme to Eliminate LF (GPELF) [4], LF remains endemic in many parts of the world. As of 2024, 485.4 million people remain vulnerable to infection [5]. To achieve elimination of LF as a public health problem, WHO recommends multiple rounds of mass drug administration (MDA), delivered annually to endemic populations [6, 7]. Since 2018, in certain settings where the standard two-drug regimen (albendazole plus diethylcarbamazine or ivermectin) has proven ineffective for reaching elimination as a public health problem, a triple-drug regimen combining all three drugs has been recommended [7].

The human-mosquito transmission cycle of the parasite allows programs to monitor progress towards elimination targets using human and/or mosquito-based indicators. For human-based indicators, this is most commonly achieved by testing blood samples for the presence of the antigen (Ag) produced by adult worms. This can be done easily using a rapid diagnostic test such as the Alere/Bioline Filariasis Test Strip (Scarborough, ME, USA) [8] or the more recently approved STANDARD Q Filariasis Ag Test (QFAT) [9, 10]. Ag testing is often combined with microscopy of dried blood slides to detect circulating Mf [11]. Recently, the use of LF antibodies as a surveillance indicator has also been explored [3, 12].

The use of molecular xenomonitoring (MX) for mosquito surveillance is becoming more common [13]. MX involves using a molecular diagnostic such as quantitative polymerase chain reaction (qPCR) to test individual or pools of mosquitoes for the presence of filarial DNA. This approach has been shown to be effective in detecting qPCR-positive mosquitoes in diverse settings such as French Polynesia [14], India [15], and American Samoa [16], as well as providing a sensitive indicator for detecting community-level changes in LF transmission in Samoa [17]. In Samoa, a study comparing an MX survey with a human Mf and Ag survey carried out at the same scale (survey time and sites covered) found that MX provided signals of transmission at more households than Mf screening [11].

WHO criteria for a country to stop MDA are based on critical cut-offs of the number of Ag-positive people detected through Transmission Assessment Surveys (TAS). Specifically, where *Aedes* mosquitoes are the main vector, critical cut-offs are calculated based on the upper boundary of the 95% confidence interval for Ag prevalence in children aged 6-7 years being below 1% [6]. These thresholds are used to guide national programs to determine when transmission has been reduced to levels low enough that it is no longer considered sustainable. Of the 16 Pacific Islands and Territories where LF is (or was) endemic, eight nations have yet to be validated as having eliminated LF as a public health problem [18]. Following numerous rounds of two-drug MDA [19], Samoa completed two rounds of triple-drug MDA in 2018 and 2023. Following the first round in 2018, which achieved a self-reported coverage of 80% [20], no significant change to Ag prevalence was observed after 6–8 months [21]. However, MX results revealed a decrease in the prevalence of infected mosquitoes at 7–9 months post-MDA [17]. The second triple-drug MDA in Samoa was delayed until September 2023 due to competing public health priorities (primarily a large measles outbreak [22] and COVID-19). By this time, the reduction in Ag prevalence, if any, that might have resulted from the first round in 2018, had been lost [21].

MX is recommended as one of the four surveillance tools for post-validation surveillance of LF by WHO in the recent guidelines for monitoring and epidemiological assessment of MDA [6]. Before this advice can be practically applied in a standardised way, there is an urgent need to fill the gaps in our knowledge about how to use this tool efficiently and understand how survey results can be interpreted in relation to human-based indicators. Additionally, the lack of defined guidelines and stop-MDA targets for MX indicators limits the use of MX for guiding programmatic decisions. Here, we report the results from two MX community surveys in Samoa conducted in 2023 and 2024 (before and after the 2023 MDA). By comparing these results to Ag and Mf prevalence in samples collected during the same time period and in the same communities, and comparing this to the previously reported Ag, Mf and MX results from 2019 [23], this study aimed to compare the impact of triple-drug MDA on both human and mosquito-based indicators in Samoa. Specifically, the objectives were:

i. To detect any evidence of ongoing LF transmission in Samoa after two rounds of triple-drug MDA distributed five years apart.
ii. To evaluate the impact of the second round of triple-drug MDA (in 2023) on human (Ag, Mf) and mosquito (filarial DNA) indicators in eight primary sampling units 10 months post-MDA.

## Methods

### Ethics

Ethics approvals were granted by the Samoa Ministry of Health and The University of Queensland Human Research Ethics Committee (protocols 2021/HE000895 and 2024/HE001263). The study was conducted in close collaboration with the Samoa Ministry of Health, the WHO country office in Samoa, and the Samoa Red Cross. Permission was sought from village leaders before entering a village. Verbal and written informed consent were obtained from all participants and from the parents or guardians of participants under the age of 18 years.

### Study region

Samoa, located in the South Pacific, is comprised of two main Islands, Upolu and Savai’i, with small populations on Manono and Apolima Islands. Samoa has a population of around 200,000 residents, with 78% residing on Upolu [24]. The country is predominately rural, with large forested areas. The majority of the population reside in small coastal villages or in the urban area around the capital Apia. Traditional housing (covered fales with no walls) is common in the rural and peri-urban areas. The primary LF vector species is *Aedes (Ae.) polynesiensis,* a highly efficient diurnal or day-biting mosquito that feeds and rests outdoors (exophilic) [25] and that can effectively maintain transmission at lower human LF infection prevalence than other mosquito species [26]. The less common night-biting *Ae. samoanus*, is also endemic [27] and serves as a secondary vector. *Culex quinquefasciatus* is also present, but considered of minor importance for LF in the Samoan Islands compared to the *Aedes* vectors [16].

### Survey design

The Surveillance and Monitoring for the Elimination of LF in Samoa (SaMELFS) project has conducted surveys before and after both the 2018 and 2023 rounds of triple-drug MDA (Fig. 1). The first two surveys occurred in 2018 [28] and 2019 [17, 23], and included 35 primary sampling units (PSUs) across Upolu, Savai’i, and Manono Island. In 2023 [21] and 2024, eight PSUs were surveyed for both human-based and mosquito-based indicators. The eight PSUs (Supplementary Fig. S1) were a subset of those surveyed in 2018 and 2019 and were chosen based on the Ag prevalence in 2019 [21]. The PSUs were selected to represent a range of transmission scenarios resulting in two each being selected from high (13-17%), medium (6-7%), low (3-5%) and zero (0%) observed Ag prevalence in 2019. All eight PSUs were located on Upolu due to logistical and cost constraints. Results from the 2018 survey have been published previously [17, 28], and not included in the current study because the human survey occurred immediately after the MDA, meaning no baseline was available for Mf. Furthermore, the 2018 MX survey included only five of the eight PSUs surveyed in 2023 and 2024.

**Fig. 1.**
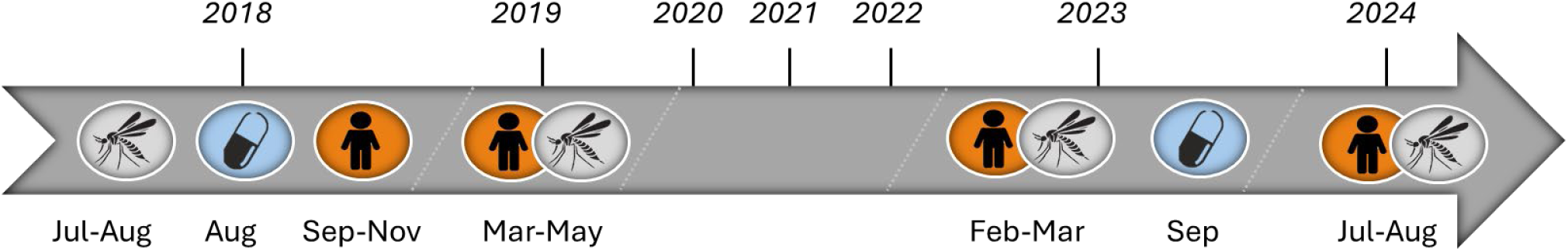
Timeline of Surveillance and Monitoring for the Elimination of Lymphatic Filariasis in Samoa (SaMELFS) human surveys (orange circles) and mosquito surveys (grey circles) in relation to the distribution of triple-drug mass drug administration (blue circles). Dates are provided in Supplementary Table S1.

### Human surveys

In each of the eight PSUs, we selected 15 houses using a virtual walk method with a random starting point [28]. Households were approached by a member of the Samoa Red Cross and offered the opportunity to participate in the survey. A household was defined as all people who shared the same kitchen. Household members were eligible to participate if they were aged ≥5 years and had slept at the property within the last week or considered it their main place of residence. Eligibility criteria were consistent over the three surveys. Target sample sizes per PSU were 57 people aged ≥10 years in 2019, 60 people aged ≥5 years in 2023 and 100 people aged ≥5 years in 2024. Specific protocols for the 2019 and 2023 surveys have been described in detail elsewhere [21, 23].

Households where all members declined participate or where no one was home at the time of the visit (or after a repeat visit if time allowed) were replaced with the nearest neighbouring household. If the target sample size had not been achieved after 15 houses had been surveyed, additional households were enrolled until the target was met. In 2019, additional the houses were selected at random. In 2023 and 2024 respectively, 5 and 10 extra random houses were preselected.

In each survey, all participants (or their guardians) were asked a series of demographic and behavioural questions, with their answers recorded electronically by bilingual field teams. A finger-prick blood sample (up to 500 µL) was collected in a heparin microtainer. The samples were kept in cooler bags before being refrigerated in the lab for one (usually) or two nights. Samples were returned to room temperature and tested for LF Ag using Alere/Bioline Filariasis Test Strips (Scarborough, ME, USA) and read at 10 minutes. Two slides of thick blood smears (three rows of 20 µL each) were made from any Ag-positive samples, following WHO guidelines [6]. In 2019 and 2023, both slides from each participant were stained and read under a microscope by two independent readers to identify and count Mf. In 2024, only one set of slides was stained, and the other was read unstained. If the Mf-positivity differed between readers, or if the difference in counts was >10%, both readers re-examined these slides to reduce the chance that observed discrepancies were due to reader error. Mf density (Mf/mL) was calculated for each Mf-positive participant as the average number of Mf counted from each of the two slides (generally 60 µL per slide) and converted to counts per mL.

### Mosquito surveys

In 2023 and 2024, mosquito surveys were conducted concurrently with the human surveys, with Biogents-Sentinel traps (BG-Sentinel) (Biogents AG, Regensburg, Germany) placed at households within two weeks of the human survey. Where possible, traps were placed at or near participant households from the human survey. BG-Sentinel traps with attractant BG-lures were placed at households for approximately 48 hours, with a mosquito collection and battery change after 24 hours. Mosquitoes were kept in a cooler box until being anaesthetised via freezing at −20 °C. Male mosquitoes were discarded and female mosquitoes were pooled by morphological species identification and trap location, with a maximum of 25 mosquitoes per pool. Samples were stored at −20 °C until being preserved. In 2019 and 2023, pooled samples were preserved by oven-drying for three hours at 60 °C. In 2024, mosquito pools were preserved in approximately 200 µL of DNA/RNA shield™ (Zymo Research, USA, Cat No. R1100-250). Full protocols for the 2023 and 2024 qPCR testing are given in Supplementary Material S1. Methods and protocols for the 2019 survey are available in McPherson et al. (2022) [17].

### Statistical analysis

We calculated Ag and Mf prevalence in each of the three surveys (2019, 2023, and 2024), overall for the eight PSUs combined, and individually for each PSU. All prevalence estimates for Ag and Mf were calculated in Stata 18.0 [29] using the ‘svyset’ command from the ‘svy’ package to account for the sampling design and probability of selection. Estimates were adjusted for age and sex distribution based on the 2016 census [30]. We also calculated the proportion of Ag-positive participants in each year who were also Mf-positive (potentially infectious). A two-sample test for equality of proportions was used to compare the difference between years. The geometric mean Mf density of Mf-positive participants was calculated for each of the three years and a two-sample t-test on the log-transformed data was used to compare the difference between 2019 and 2024, and between 2023 and 2024

All estimates of the prevalence of qPCR-positive mosquitoes were calculated in the PoolTestR package [31] using Bayesian mixed effect logistic regression models with adjustments to account for the varying pool size. In the primary analysis by genus, fixed effects for combinations of sample year and genus were used for population-level differences across all sampled sites. For each genus and year, random effects for PSU and trap site (household) were used to account for hierarchical sampling and clustering of infection by location, allowing for possible correlation between random effects for *Aedes* and *Culex* mosquitoes at each location. The degree of clustering (standard deviation of random effects) at the trap and PSU levels were estimated for each mosquito genus but assumed to be constant across sampling years. Prevalence estimates by PSU accounted for uncertainty in generalising to unsampled sites within the PSU.

Overall prevalence estimates across the eight PSUs were calculated by taking the average of estimates for each PSU, i.e., without attempting to generalise to unsampled PSUs. For each PSU, the overall prevalence in both mosquito genera combined was the average of genus-specific estimates, weighted by the number of mosquitoes of each genus tested in that PSU. Estimates for *Ae. polynesiensis* were calculated separately using a model with the same methods but with fixed effects only for year and for each year random effects by PSU and sampling site. Estimates for the prevalence of qPCR-positive mosquitoes are reported with 95% credible intervals (CrI).

To evaluate the impact of the 2023 MDA on LF infection indicators over time, we calculated the odds ratio (OR) of a positive test in 2024 compared to 2019 or 2023. We evaluated all six indicators: three human-based and three MX-based. Human-based indicators (Ag prevalence, Mf prevalence, and proportion of Ag-positive people who were Mf-positive) were calculated using Stata version 18.0 [29]. The PoolTestR package [31] was used to calculate ORs for the mosquito-based indicators (prevalence of qPCR-positive mosquitoes from all species, all *Aedes* species and *Ae. polynesiensis)*.

## Results

### Participant distribution and demographics

In 2024, there were 1,109 participants enrolled. Of these, 1,074 (97%) had valid Ag results, and 35 were missing results due to sample quantity or quality. Participant numbers and demographics for those with valid Ag results are given in Table 1. Results from 2019 and 2023 have been published previously [21, 23].

**Table 1.** Participant numbers and demographic details for the 2019, 2023, and 2024 surveys in eight sentinel primary sampling units (PSUs) in Samoa, categorised by 2019 antigen (Ag) prevalence category. Results from 2019 and 2023 have been published previously [21, 23] and are included here for comparison. Numbers include only participants with valid Ag results. PSU=primary sampling unit.

| Year |  |  | 2019 | 2023 | 2024 |
| --- | --- | --- | --- | --- | --- |
| Total participants |  |  | 643 | 623 | 1074 |
| Sex n (%) | Male |  | 291 (45.3%) | 280 (45.0%) | 494 (46.0%) |
|  | Female |  | 352 (54.7%) | 343 (55.0%) | 580 (54.0%) |
| Median age in years (range) |  |  | 20 (5-80) | 22 (5-93) | 20 (5-94) |
| Number of households |  |  | 121 | 125 | 187 |
| Number of participants (n, %) by PSU and 2019 Ag prevalence category | Zero (0%) | Vaivase Tai | 58 (9.0%) | 61 (9.8%) | 101 (9.4%) |
|  |  | Mutiatele & Salea’aumua | 89 (13.8%) | 68 (10.6%) | 132 (12.3%) |
|  | Low (3-6%) | Tuanai | 76 (11.8%) | 72 (11.6%) | 128 (11.9%) |
|  |  | Fusi | 69 (10.7%) | 91 (14.6%) | 155 (14.4%) |
|  | Medium (6-7%) | Vaiusu | 100 (15.6%) | 101 (16.2%) | 147 (13.7%) |
|  |  | Falefa | 74 (11.5%) | 58 (9.3%) | 124 (11.5%) |
|  | High (13-16%) | Faleasiu | 102 (15.9%) | 72 (11.6%) | 149 (13.9%) |
|  |  | Lauli’i | 75 (11.7%) | 100 (16.1%) | 138 (12.8%) |

### Antigen prevalence

In 2024, the overall adjusted Ag prevalence in the eight PSUs was 10.3% (95% CI:7.3-14.5) representing no significant change (p=0.94) from 2023 when it was 9.9% (95% CI: 3.5-21.0) [21] or from the 9.8% (95% CI:5.6-15.5) Ag prevalence in 2019 (Fig. 2, Supplementary Table S2). Prevalence remained highest in the known hotspots of Faleasiu and Lauli’i.

**Fig. 2.**
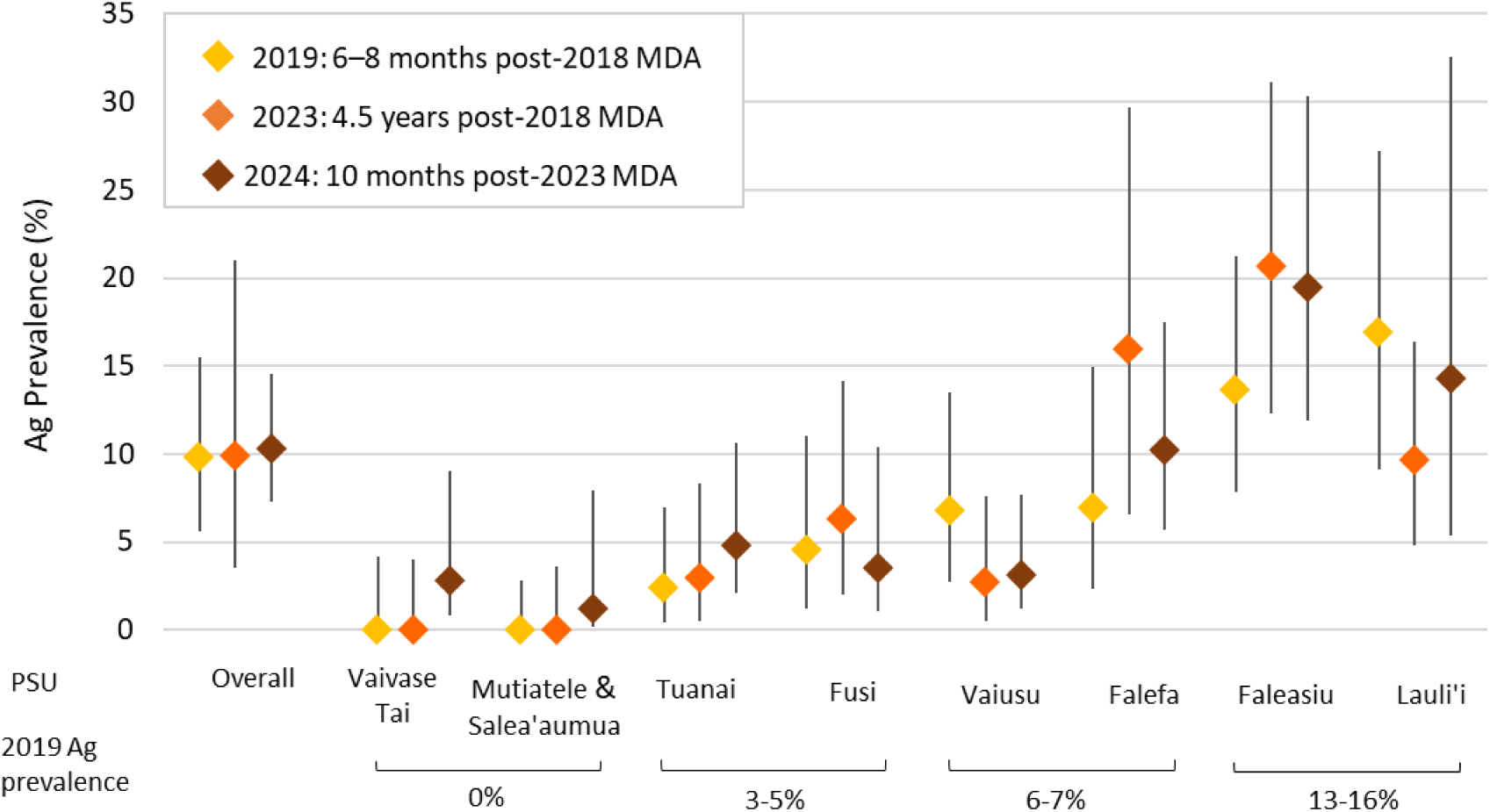
Antigen (Ag) prevalence in Samoa for the eight primary sampling units (PSUs) surveyed in 2019 (6–8 months after the 2018 mass drug administration [MDA]), 2023 (4.5 years after the 2018 MDA) and 2024 (10 months after the 2023 MDA), overall and by PSU. Error bars represent 95% CIs. The 2019 Ag prevalence categories reflect the prevalence from the 2019 survey.

### Microfilaria indicators

In 2024, the overall adjusted Mf prevalence in the eight PSUs was 3.1% (95% CI: 1.8-5.5) compared to 5.1% (95% CI: 1.3-12.4) in 2023 and 3.1% (95% CI: 1.3-5.9) in 2019 (Fig. 3, Supplementary Table S3). Geometric mean Mf density of the 19 Mf-positive participants in 2024 was 96.7 Mf/mL (range 8.3 to 6633.3). This was not statistically different to previously reported values from the 16 Mf-positive participants in 2023 (108.9 Mf/mL, range 8.3 to 800.0, p= 0.89), or the 11 Mf-positive participants from 2019 (124.2 Mf/mL, range 16.7 to 867.7, p=0.41). In 2024, 26.5% (95% CI: 16.5-38.6%) of Ag-positive participants were Mf-positive, compared to 40.1% (95% CI: 25.6-57.9%) in 2023 and 31.7% in 2019 (95% CI: 18.1-48.1).

**Fig. 3.**
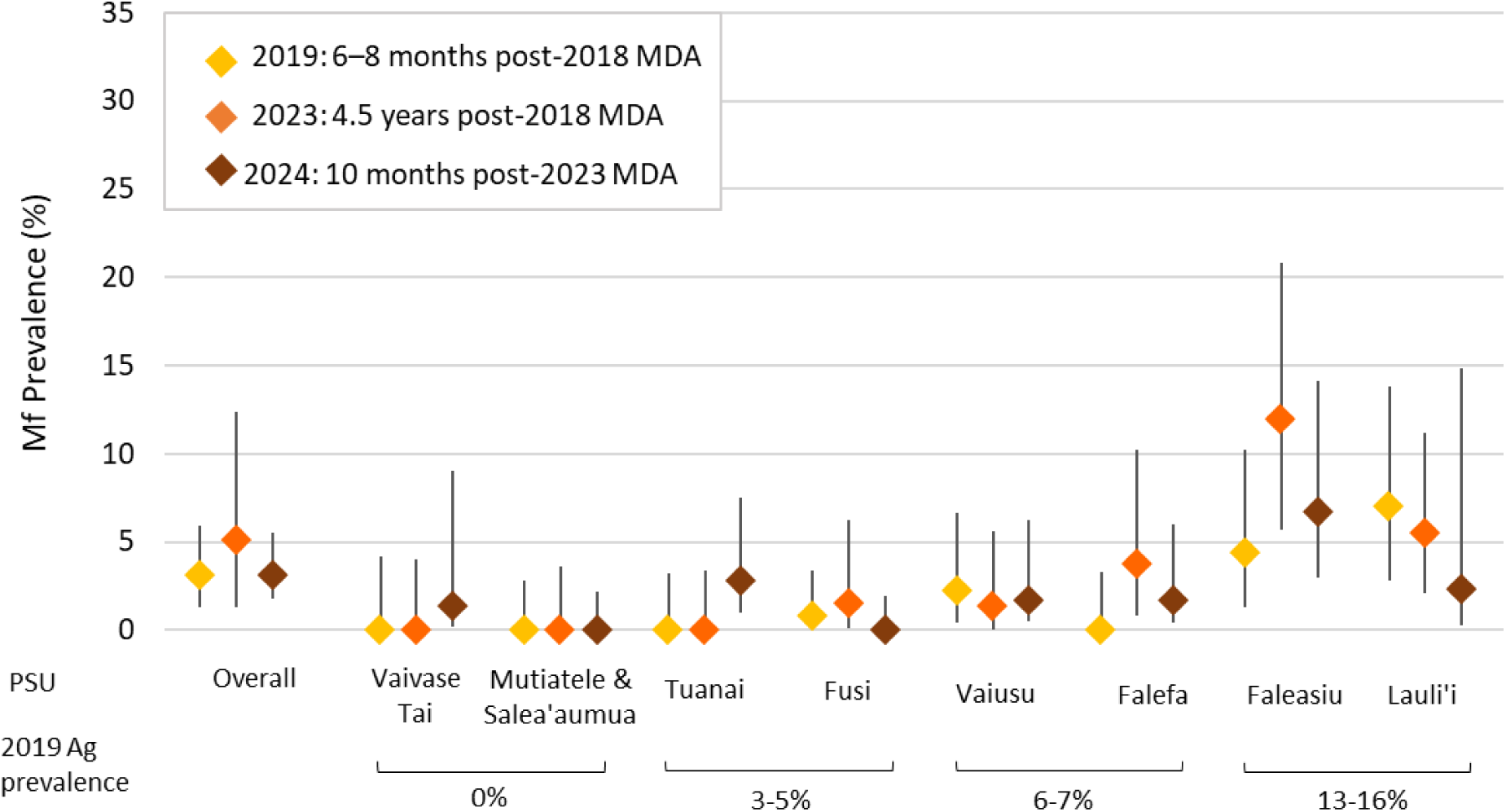
Microfilaria (Mf) prevalence in Samoa for the eight primary sampling units (PSUs) surveyed in 2019 (6–8 months after the 2018 mass drug administration [MDA]), 2023 (4.5 years after the 2018 MDA) and 2024 (10 months after the 2023 MDA), overall and by PSU. Error bars represent 95 % CIs. The 2019 Ag prevalence categories reflect the prevalence from the 2019 survey.

### Abundance and Species Diversity of Mosquitoes

In 2023, a total of 5,378 female mosquitoes were tested, grouped into 543 pools (Fig. 4, Supplementary Table S4). The average catch size per PSU was 672 (range 321 to 1127). Mean number of mosquitoes per pool was 10.9 (range 1 to 25) for *Aedes* spp. and 5.6 (range 1 to 25) for *Culex* spp. In 2024, a total of 3,914 female mosquitoes were tested, grouped into 411 pools, with a mean pool size of 10.5 (range 1 to max 25) for *Aedes* spp. and 6.8 (range 1 to 25) for *Culex* spp. (Supplementary Table S5). The average catch size per PSU was 489 (range 293 to 644).

**Fig. 4.**
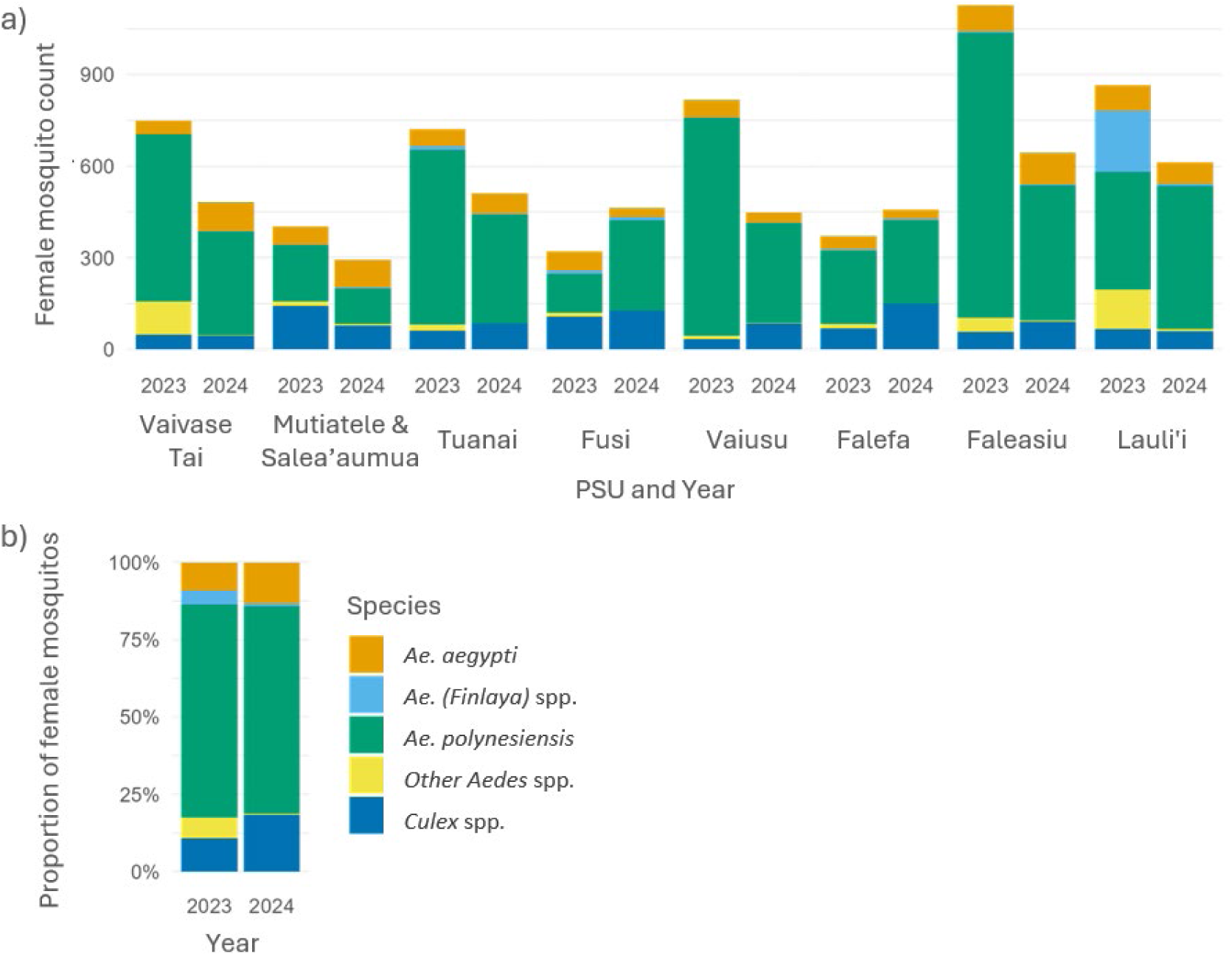
Abundance of female mosquitoes trapped in the Surveillance and Monitoring for the Elimination of LF in Samoa (SaMELFS) 2023 and 2024 MX surveys (a) counts by PSU, and (b) overall proportion by species and year.

*Ae. polynesiensis* was the most common species caught in both 2023 (3,705 mosquitoes, 69% of total catch) and 2024 (2,619 mosquitoes, 67% of total catch). *Culex* mosquitoes made up 11% of the catch in 2023 and 18% in 2024. Species abundance and diversity by PSU are presented in Fig. 4. The species distribution in 2019 has been previously reported [17] and mean pool sizes for all three years are given in Supplementary Table S5.

### Prevalence of qPCR-positive mosquitoes

In 2023, 16% (91/543) of all mosquito pools were qPCR-positive. In 2024, 8% (34 /411) of all mosquito pools were qPCR-positive. Prevalence of infected mosquitoes was consistently higher in *Aedes* spp. compared to *Culex* spp. (Fig. 5, Supplementary Table S6). Estimated prevalence by PSU are given in Supplementary Fig. S2.

**Fig. 5.**
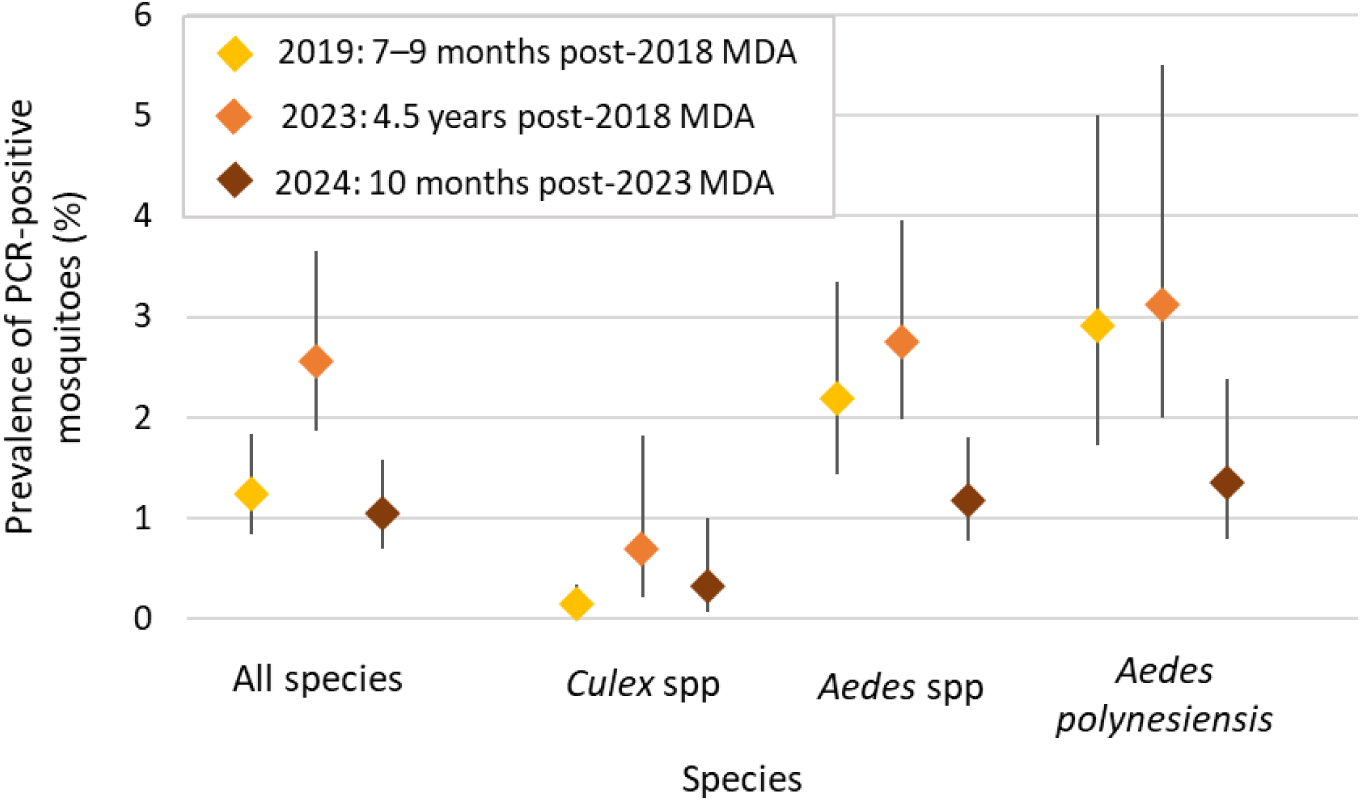
Prevalence of quantitative polymerase chain reaction (qPCR)-positive mosquitoes in eight primary sampling units in Samoa in 2019 (7–9 months after the 2018 mass drug administration [MDA]), 2023 (4.5 years after the 2018 MDA) and 2024 (10 months after the 2023 MDA) for all species, *Culex* spp., all *Aedes* spp., and *Ae. polynesiensis*. Error bars represent the 95% credible intervals.

### Impact of MDA

There was no significant change in Ag prevalence between the 2023 survey (pre-2023 MDA) and the 2024 survey, 10 months after the 2023 MDA (OR 1.0, 95% CI: 0.7, 1.6). Although there were reductions in Mf prevalence from 2023 to 2024 (OR 0.68, 95% CI:0.33-1.4) and in the proportions of Ag-positives who were Mf-positive, the differences were not statistically significant (Fig. 6, Supplementary Tables S7). For all mosquito-based indicators, ORs showed a decrease in the prevalence of qPCR-positive mosquitoes from 2023 to 2024, and also from 2019 to 2024, noting that in this second time period, the change for all species combined was not statistically significant.

**Fig. 6.**
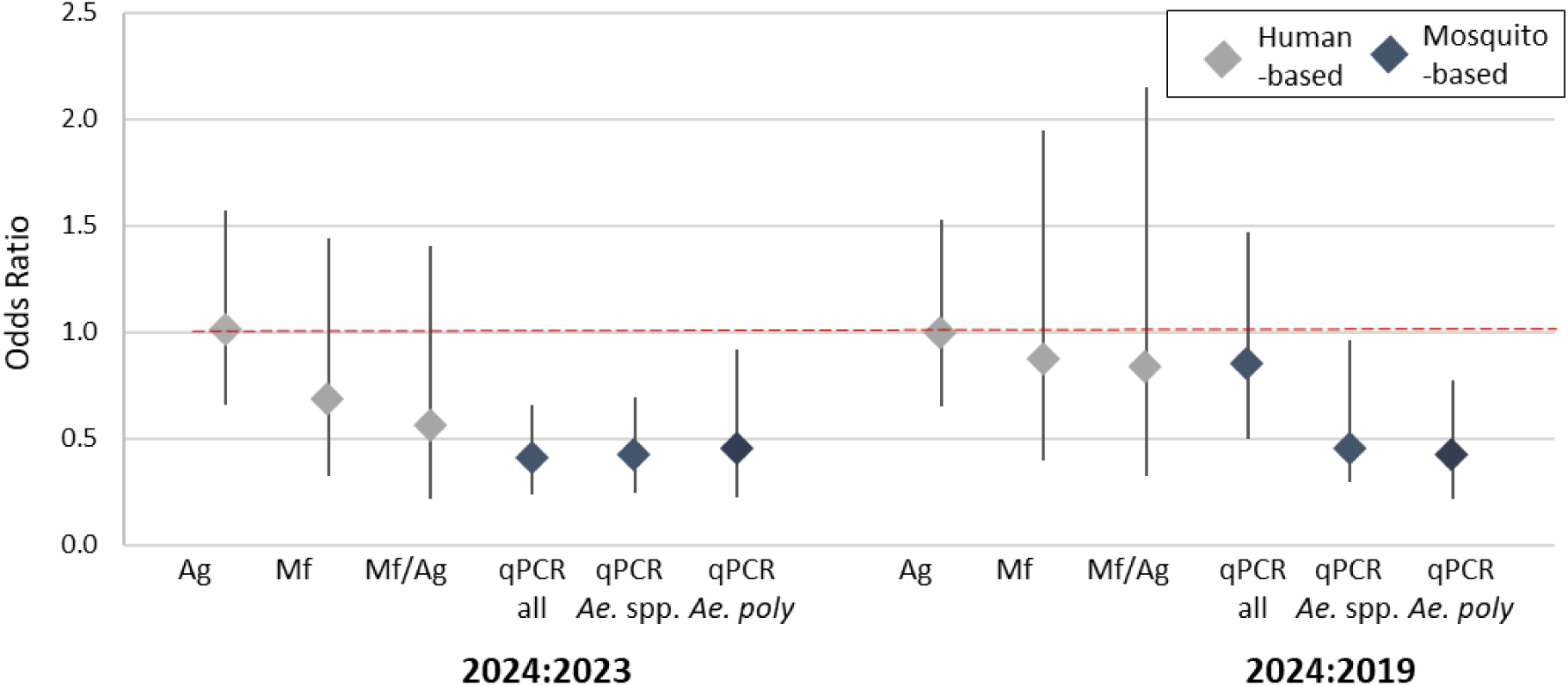
Odds ratios for human and mosquito-based LF indicators in eight primary sampling units in Samoa in 2024 (10 months post-2023 MDA) compared to 2023 (4.5 years post-2019 MDA, pre-2023 MDA), and in 2024 compared to 2019 (6–9 months post 2019-MDA). Human indicators are antigen (Ag), microfilaria (Mf), proportion of Ag-positive people who were also Mf-positive (Mf/Ag). Mosquito indicators are the positive/negative results of quantitative polymerase chain reaction (qPCR)-positive mosquitoes for all species (qPCRall), all *Aedes* species (qPCR *Ae. s*pp.) and *Aedes polynesiensis* (qPCR *Ae. poly*). Horizontal dashed line (OR = 1) indicates no change between years.

## Discussion

Two rounds of triple-drug MDA, spaced five years apart, were not sufficient to break the LF transmission cycle in Samoa. Despite no confirmed reduction in Ag or Mf prevalence following the 2023 MDA round, mosquito-based indicators detected evidence of reduced transmission 10 months after MDA. The non-significant decrease between 2023 and 2024 in both Mf prevalence and the proportion of Ag-positive participants who were Mf-positive (potentially infectious) also represents a likely decrease in transmission intensity. The timeframe needed to detect a similar signal by using Ag alone remains undetermined and it is currently unclear whether countries could expect to see a decrease in Ag prevalence prior to any recommended additional MDA rounds. While Mf prevalence showed a reduction from 2023 to 2024 (pre- and post-MDA), the sample size was insufficient to detect a statistically significant change.

Based on our previous results from Samoa [23], it was unsurprising to find that Ag prevalence had not reduced 10 months after the 2023 MDA. While a single dose of the triple-drug treatment has been shown to be effective at clearing Mf in Samoa within a week [32], Ag can persist in the blood for months or years after infection has been cleared [33]. Evidence from previous surveys in Samoa has shown that recrudescence or reinfection is also possible in those who have been treated [34, 35]. The decrease in Mf prevalence 10 months after the 2023 MDA, as well as the decrease in the proportion of Ag-positive people who are potentially infectious (Mf-positive), suggests that at a community level, at least some degree of Mf clearance is maintained over this time. However, if the reduction in Mf prevalence is not enough to reduce transmission, these gains are likely to be lost within five years [21]. It should also be considered that without the MDA, Ag prevalence may have continued to increase, meaning that we cannot be sure if the 2023 MDA slowed or halted an even greater increase in transmission, even if there was no observable decrease from current levels.

The lack of any observed change in Ag prevalence is in contrast with recent results from Papua New Guinea which showed a 30% reduction in Ag prevalence 12 months after triple-drug MDA [36]. While the contradicting results suggest a difference in MDA effectiveness between the two countries, further longitudinal data are needed to explore whether this discrepancy represents a genuine difference in the effectiveness of triple-drug MDA. The results from Papua New Guinea reported by Bun *et al.* (2025) also found a much larger decrease in Mf prevalence compared to Ag prevalence 12 months after the triple-drug MDA [36]. They concluded that Ag was not a suitable indicator for measuring current transmission in regions that have recently distributed MDA. Our results support this assertion that evaluating the impact of an MDA in the early post-intervention period (within 10 months) requires a more time-sensitive measure than Ag.

The mosquito surveillance results from this study support existing evidence from the Pacific, including Samoa [12, 18], American Samoa [17], and French Polynesia [15], that MX could strengthen efforts to monitor the impact of MDA in the region. Our results from the 2023 and 2024 surveys in Samoa confirm previous findings from 2018 and 2019 that mosquito-based indicators for filarial DNA are not only useful for providing signals of transmission at specific locations [11], but also serve as a more time-sensitive indicator than Ag for monitoring the impact of MDA in the months immediately after drug distribution [17].

In contrast to recent results from French Polynesia [14, 37] that found the highest prevalence of qPCR-positive mosquitoes in a non-vector species (*Ae. aegypti*), the highest prevalence in our study was seen in the main vector (*Ae. polynesiensis*). Prevalence of qPCR-positive mosquitoes was also consistently higher in *Aedes* spp. than in *Culex* spp. This difference was reflected in a lower overall prevalence of qPCR-positive mosquitoes in 2019 (where approximately half the samples were *Culex* spp.) compared to both 2023 and 2024, in which the majority of the catch was *Aedes* spp., raising the overall qPCR-positive prevalence.

In a context where researchers are already challenged with the analytical complexities of estimating prevalence from pooled samples, often with hierarchical survey design [13], whether further complexity should be added by combining genera presents several considerations. Our results found that sorting mosquitoes to species level (all *Aedes* spp. compared to *Ae. polynesiensis*) provided limited additional useful information for decision-makers. However, if genera were combined when comparing prevalence over time, differences in the relative number of each genus tested between timepoints can obscure trends or could create the appearance of changing prevalence where there was none. New tools that simultaneously determine the species composition of a mixed pool and test for the presence of pathogen DNA could remove the need to sort mosquitoes before pooling [38]. However, these methods are yet to be applied to large field surveys and, as of yet, there is no software available for easily conducting the appropriate statistical analysis of the resulting data. The markedly different prevalence between genera also creates a dilemma when setting targets for MX in a validation setting and raises questions as to whether separate cut-offs are required for different species and regions [13].

There are several limitations to consider when interpreting the results of this study. First, because of the limited sample size of the human surveys, the study lacked statistical power to significantly detect small year-to-year differences in Ag or Mf prevalence. In contrast, our study identified significant changes between years in the prevalence of qPCR-positive mosquitoes despite adverse weather conditions in 2024 leading to lower catch sizes of mosquitoes in some villages. Second, because the sampling design did not involve random selection of villages, overall prevalence estimates in the eight PSUs cannot be generalised to the national level and therefore cannot be considered in relation to the WHO target thresholds [6].

As endemic countries progress towards lymphatic filariasis (LF) elimination, robust monitoring and surveillance are essential to assess the impact of triple-drug mass drug administration (MDA) and inform decisions on the need for, and timing of, additional rounds [39]. Ag testing provides a core indicator for identifying populations requiring treatment but has limited sensitivity for detecting short-term changes in transmission following MDA. Mf prevalence may provide earlier evidence of impact than Ag, although its use is constrained by operational and sampling requirements. However, mosquito-based surveillance indicators have demonstrated greater temporal sensitivity to changes in transmission following MDA and can provide more timely programmatic signals than human-based indicators. Clear guidance on the programmatic use of MX is now urgently needed to support standardised survey design, analysis, and interpretation. This integrated surveillance approach, which combines human and mosquito indicators, will be critical to support timely, evidence-based programmatic decision-making to guide adaptive interventions and sustain progress towards interruption of transmission and elimination.

## Acknowledgements

We are extremely grateful to our colleagues and collaborators at Samoa Ministry of Health. This work would not have been possible without our colleagues at the Samoa Red Cross whose continued efforts supporting field surveys, especially Secretary General Namulauulu Tautala Mauala, Nixon Norman Mataia, Brenda Koon Wai You, Alesi Samuelu and Shem Lepale.

We would also like to acknowledge the significant in-country support provided by Lepaitai Hansell at the WHO office in Apia and extend our sincere thanks for her assistance during the survey preparation and fieldwork. We would like to acknowledge Filipina Amosa-Lei Sam for her assistance with translating research documents from English to Samoan. The mosquito testing protocol used in this work was based on protocols created by Richard Bradbury and his research team with assistance from Mary Doherty and Ashryella Seeler.

## Funding statement

**Funding:** This work received financial support from the Coalition for Operational Research on Neglected Tropical Diseases (COR-NTD), which is funded at The Task Force for Global Health primarily by the Bill & Melinda Gates Foundation and, previously, the United States Agency for International Development through its Neglected Tropical Diseases Programme (OPP1190754 to CLL, and G2775 to RSB, CLL, HJM, and SH). Under the grant conditions of the Foundation, a Creative Commons Attribution 4.0 Generic License has already been assigned to the Author Accepted Manuscript version that might arise from this submission. CLL was supported by an Australian National Health and Medical Research Council (NHMRC) Investigator Grant (APP1193826). This work was supported by the Operational Research and Decision Support for Infectious Diseases (ODeSI) program, which is funded by The University of Queensland’s Health Research Accelerator (HERA) initiative (2021-2028). The 2024 mosquito survey was funded by the Australian Defence Force Malaria and Infectious disease Institute (ADFMIDI). With the exception of ADFMIDI, the funders had no role in study design, data collection and analysis, decision to publish, or preparation of the manuscript.

## Data Availability

Data used in this study were collected during field surveys conducted in Samoa. Communities in Samoa are small, and sharing individual-level human data could enable identification of participants, which would violate the conditions of the study’s ethics approvals. As a result, individual-level human data are not publicly available. Requests for access to de-identified individual-level data may be made to the Human Research Ethics Committee at The University of Queensland, citing protocols 2021/HE000895 and 2024/HE001263, and will be considered subject to ethical approval and data sharing agreements. All relevant aggregated data required to interpret the findings of this study, including primary sampling unit–level estimates for human and mosquito indicators, are provided in the Supplementary Material.

## Supplementary Material 1

### Protocols for 2023 and 2024 qPCR testing of mosquito samples

**2023:**

Richard Bradbury & Asrhyella Seeler

Institute of Innovation, Science and Sustainability, Federation University, Berwik, Victoria, 3806

**2024:**

Donna MacKenzie, Lisa Rigby, Rhys Izuagbe, Jo Kizu

Australian Defence Force Malaria and Infectious Disease Institute, Enoggera, Queensland 4051, Australia

#### 2023 qPCR testing protocol

Homogenisation of mosquito tissue in each pool was performed in 1.8 mL Eppendorf tubes (Eppendorf, Hamburg, Germany), with a single 4.5 mm diameter autoclaved steel ballbearing added, in a TissueLyser (Qiagen, Hilden, Germany) at a frequency setting of 30.0 for 10 minutes, or until the macerated tissue material was homogenous. DNA was then extracted from the macerated mosquito tissue using a Qiagen DNeasy Blood and Tissue kit (Qiagen, Hilden, Germany) according to the manufacturer’s protocol. A PCR grade water negative control was incorporated into each series of extractions performed.

qPCR was performed on mosquito DNA extracts using the Wb-Cl1 qPCR protocol provided by Quinnipiac University − Pilotte Lab [1]. All qPCR assays were performed using a Realplex 2 Mastercycler (Eppendorf, Hamburg). Each assay consisted of 12.5 µL of PrimeTime® Gene Expression Master Mix (Integrated DNA Technologies, Coraville, IA), 1 µM of forward and reverse primer, 250 nM of TaqMan probe, 6.25 µL of PCR grade water (Integrated DNA Technologies, Coraville, IA) and 5 µL of sample DNA extract. A DNA extract from a previously collected known positive W. bancrofti blood sample and a blank DNA extraction of PCR-grade water were included with each run as positive and negative controls, respectively. Any amplification ≤40 cycles was considered a positive result. Discordant results were repeated a third time, with the consensus of the three assays chosen as the valid result.

[1] Zulch, M.F., N. Pilotte, J.R. Grant, C. Minetti, L.J. Reimer, and S.A. Williams, Selection and exploitation of prevalent, tandemly repeated genomic targets for improved real-time PCR-based detection of Wuchereria bancrofti and Plasmodium falciparum in mosquitoes. PLOS ONE, 2020. **15**(5): p. e0232325.

#### 2024 qPCR testing protocol

Homogenisation of mosquito tissue in each pool was performed on a TissueLyser (Qiagen, Hilden, Germany, Cat No. 85600) for 4 minutes at 50 oscillations per second. Pooled mosquito tissue was lysed using the QIAamp® DNA Mini Kit (Qiagen, Cat. No. 51306) according to the manufacturer’s instructions. DNA extraction was performed using MagMAX™ Prime Viral/Pathogen Nucleic Acid Isolation Kit (Thermo Fisher Scientific, Waltham, Massachusetts, USA, Cat no. A58145) on an Applied Biosystems MagMax™ Express 96 Magnetic particle processor 710. The total sample lysate (approximately 600 µL) was aliquoted into a deep well 96-sample plate (Thermo Fisher Scientific, Cat no. 950405450). A binding bead mix (285 µL of Binding Solution and 10 µL Total Nucleic Acid Binding Beads per reaction) was added to each sample in the deep well sample plate. The prepared deep well plate was loaded onto the KingFisher™ Flex Purification System and extraction was performed using MagMax_MVP_Standard protocol. Elutions (50 µL) were collected into a KingFisher™ 96-well microplate (Thermo Fisher Scientific, Cat no. 97002540) and sealed with MicroAmp™ Optical Adhesive Film (Applied Biosystems, Thermo Fisher Scientific).

Real-time PCR (qPCR) was performed on mosquito DNA extracts using the Wb-Cl1 qPCR protocol provided by Quinnipiac University − Pilotte Lab. For 50 reactions, 15.6 µL of 10 μM forward primer (GCTGAAAAACATTCGCTTTTGAATG), 10 μM reverse primer (GGGTAATTAAACCGGTGATCT) and 31 µL of 10 μM double quenched probe (56-FAM/ ACAACAACT/ZEN/ATATGGGAATGGTGCAGGT/3IABKFQ, Integrated DNA Technologies, Coraville, IA) were added to 625 µL of TaqPath™ Pro Amp™ Master Mix (Applied Biosystems™ Cat A30865) and 312.8 µL of nuclease free water. Each reaction contained 5 µL of DNA template.

Samples were run on a Bio Molecular Systems Magnetic Induction Cycler (MIC) PCR (Bio Molecular Systems, Queensland, Australia) under the following cycling conditions: 50°C for 2 minutes, 95°C for 10 minutes, followed by 40 cycles of 95°C for 15 seconds and 60°C for 1 minute. Samples were classified as qPCR positive if there was amplification within 39 cycles (i.e., C_t_ <39). Initial qPCR screened positive samples were re-screened in duplicate. Data was analysed on MIC qPCR software v2.10.0.

**Supplementary Fig. S1.**
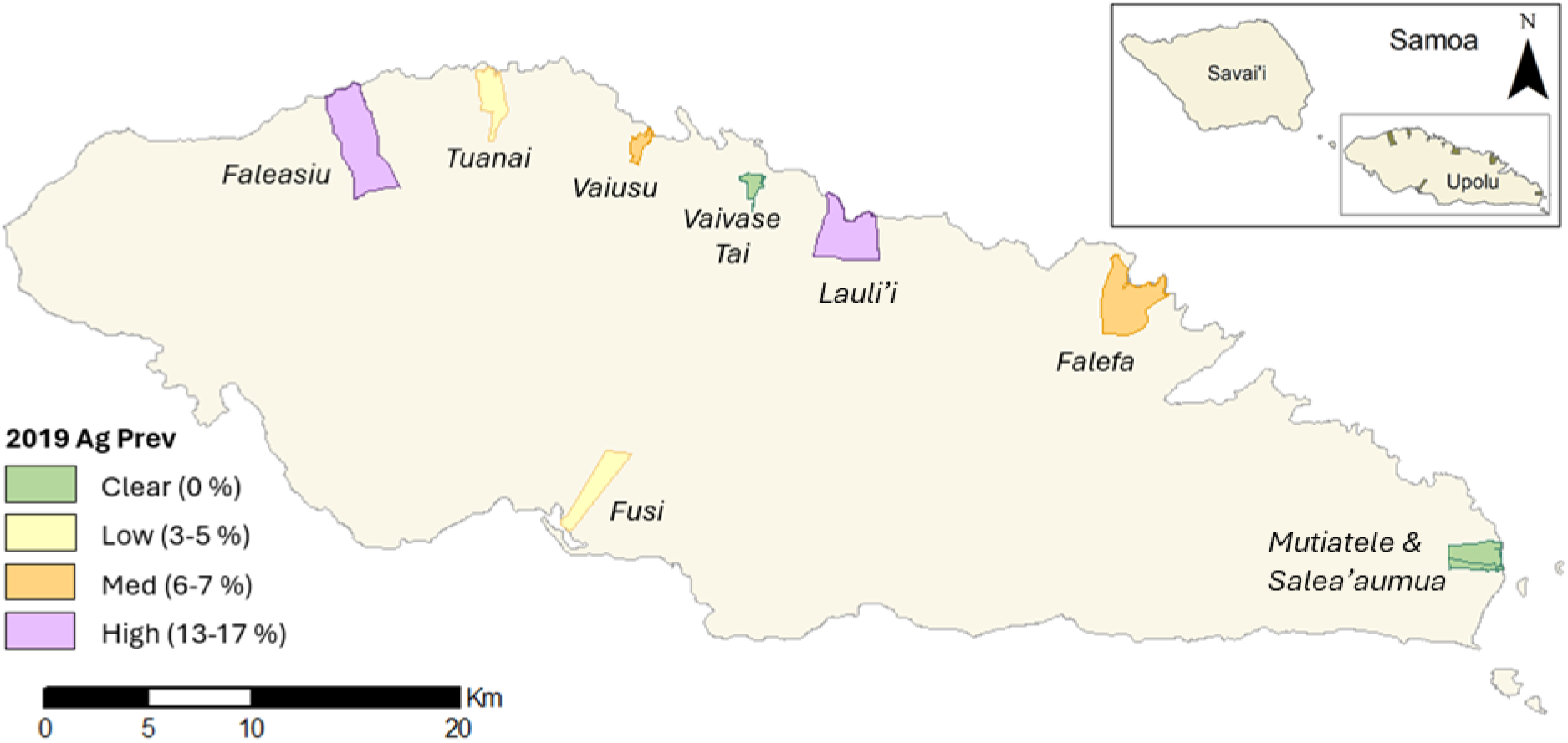
Selected primary sampling units on the Island of Upolu in Samoa which were surveyed in each of the 2019, 2023, and 2024 as part of the Surveillance and Monitoring for the Elimination of LF in Samoa (SaMELFS) program. Colour indicates the estimated antigen prevalence at baseline (2019). Spatial data on country, island, region, and village boundaries in Samoa were obtained from the Pacific Data Hub (pacificdata.org accessed on 8 July 2020) and DIVA-GIS (diva-gis.org, accessed on 12 August 2019) under an open access licence available at https://pacific-data.sprep.org/resource/public-data-license-agreement-0.

**Supplementary Fig. S2.**
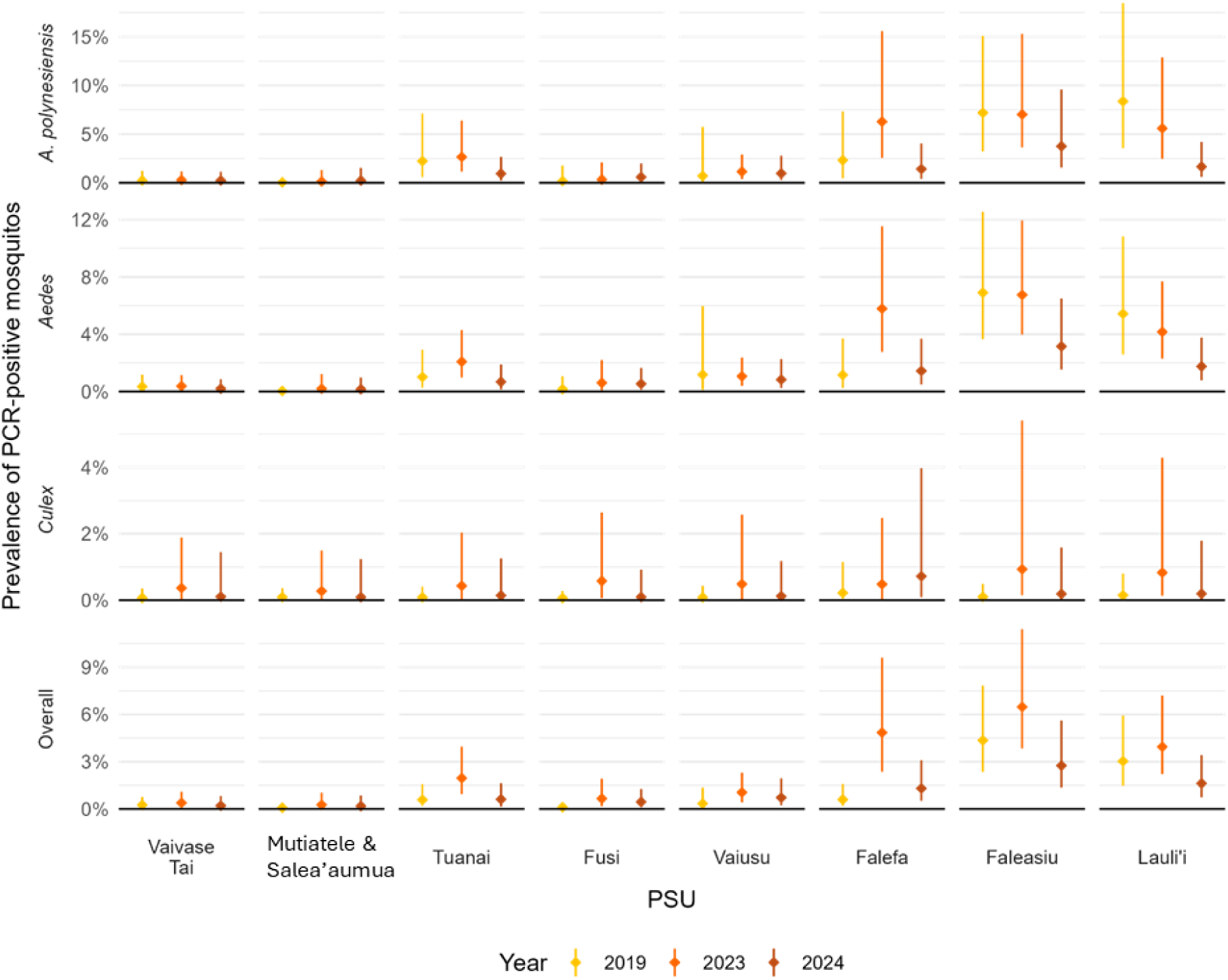
PSU-level prevalence of qPCR-positive mosquitoes in the eight primary sampling units in the Surveillance and Monitoring for the Elimination of LF in Samoa (SaMELFS) program surveys in 2019, 2023, and 2024. Note that y-axes differ by row.

**Supplementary Table S1.** Dates for the Surveillance and Monitoring for the Elimination of LF in Samoa (SaMELFS) program surveys. Triple-drug mass drug administration was implemented by Samoa Ministry of Health from the 14th to the 26^th^ of August 2018 and 16^th^ to 24th September 2023.

| Year | Activity | Start date | End date |
| --- | --- | --- | --- |
| 2018 | Mosquito survey | 2 Jul | 25 Aug |
| 2018 | Human survey | 26 Sep | 09 Nov |
| 2019 | Human survey | 28 Mar | 17 May |
| 2019 | Mosquito survey | 20 May | 6 Jul |
| 2023 | Human survey | 2 Mar to | 15 Mar |
| 2023 | Mosquito survey | 8 Mar | 20 Mar |
| 2024 | Human survey | 25 Jul | 17 Aug |
| 2024 | Mosquito survey | 17 Jul | 29 Jul |

**Supplementary Table S2.** Antigen (Ag) prevalence (%) in Samoa for the eight primary sampling units (PSUs) surveyed in 2019 (6**–**8 months after the 2018 MDA), 2023 (4.5 years after the 2018 MDA), and 2024 (10 months after the 2023 MDA).

| **Primary**<br>**Sampling**<br>**Unit** | **2019**<br>**(95% CI)** | **2023**<br>**(95% CI)** | **2024**<br>**(95% CI)** |
| --- | --- | --- | --- |
| **Overall** | **9.8**<br>**(5.6-15.5)** | **9.9**<br>**(3.5-21.0)** | **10.3**<br>**(7.3-14.5)** |
| Vaivase Tai | 0.0<br>(0.0-4.2) | 0.0<br>(0.0-4.0) | 2.8<br>(0.8-9.2) |
| Mutiatele &<br>Salea'aumua | 0.0<br>(0.0-2.8) | 0.0<br>(0.0-3.6) | 1.2<br>(0.2-7.9) |
| Tuanai | 2.4<br>(0.4-7.0) | 2.9<br>(0.5-8.3) | 4.8<br>(2.1-10.6) |
| Fusi | 4.5<br>(1.2-11.0) | 6.3<br>(2.0-14.1) | 3.5<br>(1.1-10.4) |
| Vaiusu | 6.8<br>(2.7-13.5) | 2.7<br>(0.5-7.6) | 3.1<br>(1.2-7.7) |
| Falefa | 6.9<br>(2.3-14.9) | 15.9<br>(6.6-29.7) | 10.2<br>(5.7-17.5) |
| Faleasiu | 13.6<br>(7.8-21.2) | 20.6<br>(12.3-31.1) | 19.5<br>(11.9-30.3) |
| Lauli'i | 16.9<br>(9.1-27.2) | 9.6<br>(4.8-16.4) | 14.3<br>(5.4-32.6) |

**Supplementary Table S3.** Microfilaria (Mf) prevalence (%) in Samoa for the eight primary sampling units (PSUs) surveyed in 2019 (6–8 months after the 2018 MDA), 2023 (4.5 years after the 2018 MDA), and 2024 (10 months after the 2023 MDA), overall and by PSU.

| **Primary**<br>**Sampling**<br>**Unit** | **2019**<br>**(95% CI)** | **2023**<br>**(95% CI)** | **2024**<br>**(95% CI)** |
| --- | --- | --- | --- |
| **Overall** | **3.1**<br>**(1.3-5.9)** | **5.1**<br>**(1.3-12.4)** | **3.1**<br>**(1.8-5.5)** |
| Vaivase Tai | 0.0<br>(0.0-4.2) | 0.0<br>(0.0-4.0) | 1.4<br>(0.2-9.0) |
| Mutiatele &<br>Salea'aumua | 0.0<br>(0.0-2.8) | 0.0<br>(0.0-3.6) | 0.0<br>(0.0-2.2) |
| Tuanai | 0.0<br>(0.0-3.2) | 0.0<br>(0.0-3.4) | 2.8<br>(1.0-7.5) |
| Fusi | 0.8<br>(0.1-3.4) | 1.5<br>(0.1-6.2) | 0.0<br>(0.0-1.9) |
| Vaiusu | 2.2<br>(0.4-6.6) | 1.3<br>(0-5.6) | 1.7<br>(0.5-6.2) |
| Falefa | 0.0<br>(0.0-3.3) | 3.7<br>(0.8-10.2) | 1.7<br>(0.4-6.0) |
| Faleasiu | 4.4<br>(1.3-10.2) | 11.9<br>(5.7-20.8) | 6.7<br>(3.0-14.1) |
| Lauli'i | 7.0<br>(2.8-13.8) | 5.5<br>(2.1-11.2) | 2.3<br>(0.3-14.8) |

**Supplementary Table S4.** Counts of female mosquitoes trapped by primary sampling units (PSU) the Surveillance and Monitoring for the Elimination of LF in Samoa (SaMELFS) program surveys in 2019, 2023, and 2024. See Fig. 4 in main paper for a visualisation of counts in 2023 and 2024. * This category is included in ‘Other *Aedes*’ in Figure 3.

| Year | Species | Primary Sampling Unit |  |  |  |  |  |  |  |
| --- | --- | --- | --- | --- | --- | --- | --- | --- | --- |
|  |  | Faleasiu | Falefa | Fusi | Lauli'i | Salea'au<br>mua &<br>Mutiatele | Tuanai | Vaiusu | Vaivase<br>Tai |
| 2019 | *Ae. aegypti* | 77 | 71 | 154 | 61 | 131 | 149 | 34 | 161 |
|  | *Ae. (Finlaya) spp* | 4 | 10 | 15 | 34 | 32 | 1 | 4 | 1 |
|  | *Ae. polynesiensis* | 446 | 146 | 116 | 255 | 683 | 164 | 22 | 362 |
|  | Other *Aedes* | 21 | 4 | 2 | 17 | 268 | 28 | - | 12 |
|  | *C. quinquefasciatus* | 332 | 457 | 763 | 314 | 928 | 314 | 230 | 327 |
|  | Other | 3 | - | - | 1 | 1 | 1 | - | 1 |
| 2023 | *Ae. aegypti* | 85 | 43 | 63 | 83 | 58 | 55 | 57 | 44 |
|  | *Ae. (Finlaya) spp* | 4 | 5 | 10 | 202 | 3 | 14 | 3 | 1 |
|  | *Ae. polynesiensis* | 935 | 241 | 127 | 385 | 184 | 571 | 714 | 548 |
|  | *Ae. upolensis* * | - | 2 | - | - | 3 | - | - | - |
|  | Other *Aedes* | 46 | 13 | 14 | 130 | 11 | 20 | 11 | 108 |
|  | *C. quinquefasciatus*† | 56 | 67 | 107 | 66 | 140 | 61 | 33 | 49 |
|  | Other *Culex*† | 1 | 1 | - | - | 3 | 1 | - | - |
| 2024 | *Ae. aegypti* | 103 | 28 | 31 | 70 | 89 | 67 | 34 | 95 |
|  | *Ae. (Finlaya) spp* | 3 | 7 | 11 | 8 | 5 | 3 | 2 | 1 |
|  | *Ae. polynesiensis* | 444 | 273 | 294 | 469 | 116 | 357 | 327 | 339 |
|  | Other *Aedes* | 3 | - | 1 | 7 | 6 | - | 1 | 2 |
|  | *C. quinquefasciatus*† | 91 | 149 | 127 | 59 | 77 | 85 | 85 | 45 |

† These categories are collapsed into a single category ‘*Culex*’ in Figure 3.

**Supplementary Table S5.** Mean (min-max) number of female mosquitoes per pool in the Surveillance and Monitoring for the Elimination of LF in Samoa (SaMELFS) program surveys in 2023 and 2024.

| Primary Sampling Unit | *Aedes* |  | *Culex* |  |
| --- | --- | --- | --- | --- |
|  | 2023 | 2024 | 2023 | 2024 |
| Overall | 10.9 (1–25) | 10.5 (1–25) | 5.6 (1–25) | 6.8 (1–25) |
| Faleasiu | 14.3 (1–25) | 11.5 (1–25) | 4.4 (1–11) | 7.6 (1–25) |
| Falefa | 7.2 (1–25) | 9.6 (1–25) | 6.8 (1–25) | 9.9 (1–25) |
| Fusi | 5.2 (1–21) | 9.9 (1–25) | 7.1 (1–20) | 7.9 (1–24) |
| Lauli'i | 11.4 (1–25) | 11.8 (1–25) | 5.1 (1–20) | 5.4 (1–23) |
| Salea'aumua & Mutiatele | 6.6 (1–25) | 6.4 (1–19) | 9.5 (2–25) | 5.9 (1–14) |
| Tuanai | 11.2 (1–25) | 11.5 (1–25) | 4.1 (1–13) | 6.1 (1–25) |
| Vaiusu | 13.5 (1–25) | 10.1 (1–25) | 3 (1–11) | 6.5 (1–20) |
| Vaivase Tai | 13 (1–25) | 11.8 (1–25) | 3.8 (1–10) | 3.8 (1–12) |

**Supplementary Table S6.** Prevalence of qPCR-positive mosquitoes in eight PSUs in Samoa in 2019 (7–9 months after the 2018 MDA), 2023 (4.5 years after the 2018 MDA) and 2024 (10 months after the 2023 MDA) for all species, *Culex* spp., all *Aedes* spp., and *Ae. polynesiensis*.

| Species | 2019<br>(95% CI) | 2023<br>(95% CI) | 2024<br>(95% CI) |
| --- | --- | --- | --- |
| All species | 1.2<br>(0.8-1.8) | 2.5<br>(1.9-3.7) | 1.1<br>(0.7-1.6) |
| *Culex* spp. | 0.1<br>(0.0-0.3) | 0.7<br>(0.2-1.8) | 0.3<br>(0.1-1.0) |
| *Aedes* spp. | 2.2<br>(1.4-3.3) | 2.7<br>(2.0-4.0) | 1.2<br>(0.8-1.8) |
| *Aedes polynesiensis* | 2.9<br>(1.7-5.0) | 3.1<br>(2.0-5.5) | 1.3<br>(0.8-2.4) |

**Supplementary Table S7.** Odds ratios for human and mosquito-based LF indicators in eight primary sampling units in Samoa in 2024 (10 months post-2023 MDA) compared to 2023 (4.5 years post-2019 MDA, pre-2023 MDA), and in 2024 compared to 2019 (6–9 months post 2019-MDA). Human indicators are antigen, microfilaria, proportion of Antigen-positive people who were also Microfilaria-positive.

| Survey | Indicator | Timeframe |  |
| --- | --- | --- | --- |
|  |  | 2024-2019 | 2024-2023 |
| Human | Antigen | 0.99 (0.65-1.52) | 1.01 (0.66-1.56) |
|  | Microfilaria | 0.87 (0.41-1.94) | 0.68 (0.33-1.43) |
|  | Microfilaria-positive/<br>Antigen positive | 0.84 (0.33-2.14) | 0.56 (0.22-1.39) |
| Mosquito | qPCR All species | 0.85 (0.50-1.46) | 0.41 (0.24-0.65) |
|  | qPCR *Aedes* spp. | 0.45 (0.30-0.95) | 0.42 (0.25-0.69) |
|  | qPCR *Aedes polynesiensis* | 0.42 (0.22-0.77) | 0.45 (0.23-0.91) |

## Notes

### Competing Interest Statement

The authors have declared no competing interest.

